# Simple Models Versus Deep Learning in Detecting Low Ejection Fraction From The Electrocardiogram

**DOI:** 10.1101/2024.02.06.24302412

**Authors:** J. Weston Hughes, Sulaiman Somani, Pierre Elias, James Tooley, Albert J. Rogers, Timothy Poterucha, Christopher M. Haggerty, David Ouyang, Euan Ashley, James Zou, Marco V. Perez

## Abstract

**Importance:** Deep learning methods have recently gained success in detecting left ventricular systolic dysfunction (LVSD) from electrocardiogram waveforms. Despite their impressive accuracy, they are difficult to interpret and deploy broadly in the clinical setting.

**Objective:** To determine whether simpler models based on standard electrocardiogram measurements could detect LVSD with similar accuracy to deep learning models.

**Design:** Using an observational dataset of 40,994 matched 12-lead electrocardiograms (ECGs) and transthoracic echocardiograms, we trained a range of models with increasing complexity to detect LVSD based on ECG waveforms and derived measurements. We additionally evaluated models in two independent cohorts from different medical centers, vendors, and countries.

**Setting:** The training data was acquired from Stanford University Medical Center. External validation data was acquired from Cedars-Sinai Medical Center and the UK Biobank.

**Exposures:** The performance of models based on ECG waveforms in their detection of LVSD, as defined by ejection fraction below 35%.

**Main outcomes:** The performance of the models as measured by area under the receiver operator characteristic curve (AUC) and other measures of classification accuracy.

**Results:** The Stanford dataset consisted of 40,994 matched ECGs and echocardiograms, the test set having an average age of 62.13 (17.61) and 55.20% Male patients, of which 9.72% had LVSD. We found that a random forest model using 555 discrete, automated measurements achieves an area under the receiver operator characteristic curve (AUC) of 0.92 (0.91-0.93), similar to a deep learning waveform model with an AUC of 0.94 (0.93-0.94). Furthermore, a linear model based on 5 measurements achieves high performance (AUC of 0.86 (0.85-0.87)), close to a deep learning model and better than NT-proBNP (0.77 (0.74-0.79)). Finally, we find that simpler models generalize better to other sites, with experiments at two independent, external sites.

**Conclusion:** Our study demonstrates the value of simple electrocardiographic models which perform nearly as well as deep learning models while being much easier to implement and interpret.

## Introduction

Left ventricular systolic dysfunction (LVSD) is a characteristic feature of patients with heart failure, historically with limited options for screening^1^. NT-proBNP, a laboratory biomarker that has been proposed for heart failure screening, is inexpensive, but has demonstrated only modest performance^2,3^. Transthoracic echocardiogram (TTE) screening provides a gold-standard diagnosis, but is expensive and time-consuming^4^. An ideal screening tool would be inexpensive and use information already available during routine care while offering high accuracy. One candidate is the electrocardiogram (ECG), an inexpensive and common diagnostic tool. Historically, no reliable methods existed to diagnose LVSD as defined by reduced ejection fraction from the ECG^5^, but recently deep learning methods based on the ECG waveform have demonstrated promising performance^6^, spurring multiple clinical trials^7–9^. This same trend is true for other important tasks including detecting atrial fibrillation in sinus rhythm^10^, detecting valvular disease^11^, and predicting future mortality^12,13^, leading to a great deal of excitement around the deployment of deep learning models for electrocardiography^14^.

While highly performant, deep learning has several key limitations. Domain shifts, which are difficult to track in complex distributions such as waveforms and can occur such as when a model is applied at a new hospital^15^, population^16,17^, or imaging vendor^18^, can degrade model performance significantly. Spurious correlations can allow the model to “cheat” without learning clinically salient features, for example by detecting the presence of a pacemaker or laterality marker in a chest x-ray^19,20^ or a surgical skin marking in a dermatology image^21^, leading to unintended shifts in performance during deployment. The black-box nature of neural networks can make it more difficult to interrogate them to understand the mechanisms they rely on, and attempts to remedy this issue have for the most part fallen short^22^. Technical challenges around electronic health record system integration, institutional resistance, and regulatory issues around acting on raw signals can also make model deployment prohibitively difficult^23–25^. These limitations continue to motivate the development of simpler, more interpretable methods over deep learning^26^.

Whether simpler ECG-based methods can offer similar performance to deep learning methods on complex tasks such as detecting LVSD remains unclear. Historically, diagnostic criteria to identify less complex conditions from the ECG have taken the form of simple criteria^27,28^, decision trees^29^, and linear models^30^ based on simple, hand-measurable features like the PR interval and the amplitude of the T wave. In these simpler cases, deep learning offers only slight gains over human over-reading^31^. There are multiple recent efforts to create these simple rules in a data-driven way: recently a large dataset of such measurements was mined to build a simple model to detect atrial fibrillation in sinus rhythm, though this model far under-performed deep learning methods^10,33^, and at least one other deep learning model was presented alongside a strong decision tree-based baseline based on automated measurements^12^.

In this study, we set out to understand how simpler models based on automated ECG measurements compare to deep learning models in detecting LVSD from the ECG. Using a dataset of matched 12-lead ECGs and TTEs from Stanford University Medical Center, we trained a range of models with increasing complexity to detect low ejection fraction based on ECG waveforms and derived measurements. We found that a random forest model on 555 discrete, automated measurements performs similarly to deep learning methods, and a linear model based on 5 automated measurements performs only slightly worse than deep learning but much better than NT-proBNP. This continuum of models, trading off complexity and accuracy, demonstrates that simpler methods can sometimes be substituted for deep learning models based on derived measurements, allowing for greater interpretability and ease of implementation.

## Methods

### Study populations and data sources

We trained and primarily evaluated a range of simple-measurement based and deep learning models to detect left ventricular systolic dysfunction (LVSD), as defined as a left ventricular ejection fraction below 35%, from electrocardiograms (ECG). Models were trained using a dataset of paired 12-lead resting ECGs and transthoracic echocardiograms (TTEs) from Stanford University Medical Center. This dataset consisted of all TTEs that were taken during the course of clinical care between March 2008 to May 2018 with an ECG within two weeks. ECGs that did not pass the Phillips TraceMaster quality control were removed. We extracted 39,019 TTE-ECG pairs from 27,763 patients, which were then split by patient into train, validation, and test sets in a 5:1:4 ratio. In the test set, we only included the first ECG per patient (Figure 2). These ECGs were saved as 10 second signals from all 12 leads of the ECG, sampled at 500Hz. We extracted ECG waveforms at 250Hz, along with measurements and text overreads from TraceMaster. We included all 555 measurements which had numerical values pertaining to waveform structure.

Left ventricular ejection fractions (LVEF) were extracted from STARR-OMOP^34^, a common data model of Stanford electronic health records, based on echocardiograms acquired using iE33, Sonos, Acuson SC2000, Epiq 5G or Epiq 7C ultrasound machines and interpreted by cardiologist during standard clinical practice. We included all measurements within two weeks of a record of an echo procedure. We defined LVSD as LVEF below 35%. We also extracted NT-proBNP from STARR-OMOP and included all records within 30 days of the reference ECG.

A dataset from Columbia Irving Medical Center was used as a first external validation cohort. The Columbia dataset was constructed similarly to Stanford’s, but a different ECG vendor (the General Electric MUSE system) was utilized. After inclusion criteria were applied, a random subsample of data was included for analysis. We additionally used a second external dataset from the UK Biobank. This cohort is substantially different from the first two hospital-based datasets, being made up of a cross-section of mostly healthy British patients. In the UK Biobank, all patients with a 12-lead resting ECG and cardiac magnetic resonance imaging (cMRI) taken at the first imaging visit were included. All paired ECG and cMRI studies took place on the same day; details of the cMRI protocol are available in the literature^35^. A previously described deep learning pipeline^36^ was used to estimate left ventricular ejection fraction from the cMRI. Other ECG abnormalities were determined based on the ECG text overread using string matching, validated by manual inspection. The UK Biobank ECGs were also recorded using General Electric ECG machines.

### Model development and training

Deep learning models were trained using Python 3.9 and PyTorch 1.11 on single Nvidia Titan Xp GPUs using Stanford’s Sherlock computing cluster. We closely followed the architecture described in previous literature for detecting LVSD^6^, and found that exploring different architectures did not provide a significant increase in validation AUROC. To evaluate deep learning models at other sites, we ran the model on data using a range of pre-processing parameters (with and without band pass filters, wandering baseline filters, and per-lead normalization) and reported the best performance, since different sites and vendors may use different preprocessing and follow different distributions. Random forest models were trained using Python 3.9 and XGBoost 1.7, using the binary logistic loss. We trained several models with different tree depths and numbers of trees using grid search, and selected the best model based on validation AUROC. Linear models were trained using Python 3.9 and Scikit-Learn 1.2 using standard logistic regression, without regularization or normalization unless otherwise mentioned. All analyses were performed by training models on the training set and selecting variables, hyperparameters, and models based on results in the validation set. After models were finalized, performance was evaluated on the test set and external validation sets. To select a shortlist of variables for smaller models, we selected a list of variables familiar to clinicians based on inspection and iteratively fit increasingly lasso-regularized models while removing correlated variables. We selected a model of a size where removing any one variable would cause a drop in performance of greater than 1%, while adding any one variable would cause an increase in performance less than 1%.

### Statistical Analysis

We primarily compared models based on the area under the receiver operator characteristic curve (AUROC), a standard metric used for evaluating a predictor’s performance across multiple cutoffs in binary classification tasks. All AUROCs were computed using the scikit-learn Python package. We additionally compute sensitivity, specificity, and positive predictive values using standard definitions. We report balanced sensitivity and specificity (choosing the cutoff which minimizes the difference between sensitivity and specificity), positive predictive value at the same cutoff, sensitivity at 90% specificity, and specificity at 90% sensitivity. All confidence intervals are 95% intervals generated through bootstrapping with 1,000 samples.

## Results

### Study Population

We trained several models on ECGs paired with TTEs from Stanford University Medical Center taken between March 2008 and May 2018 during the normal course of clinical practice (Figure 2). From the 96,361 resting TTEs (from 54,045 patients) with a recorded ejection fraction, 46,254 (32,361 patients) occurred within two weeks of a unique ECG. Among those, 40,994 ECGs (28,949 patients) passed the automated quality control test performed by the Philips TraceMaster software. We randomized those pairs by patient 50%/10%/40% into train, validation, and test sets, resulting in 20,269 training ECG-TTE pairs (14,448 patients), 4,276 validation ECG-TTE pairs (2,983 patients), and 16,449 ECG-TTE pairs (11,518 patients) randomized to the test group, of which 11,518 first ECGs per patient were included in the test set. In the train, validation, and test sets, 2,175 (10.73%), 462 (10.80%), and 1,119 (9.72%) ECGs, respectively, were taken from patients with LVSD (in the test set, this was also the number of patients). Detailed demographic data are shown in Table 1.

**Table 1.** Demographics in each split. Blank entries are missing: hispanic ethnicity was not tracked in the UK Biobank, race and ethnicity were not tracked for all patients in the Columbia cohort, and ECG findings were not available in the Columbia cohort.

|  | Train | Valid | Test | UK Biobank | Columbia |
| --- | --- | --- | --- | --- | --- |
| LVSD | 2,175<br>(10.73%) | 462 (10.80%) | 1,119 (9.72%) | 96 (0.28%) | 4,656<br>(12.59%) |
| LVEF | 55.35 (13.31) | 55.36 (13.36) | 56.25 (13.21) | 59.57 (6.16) | 53.85 (13.49) |
| Age | 61.46 (18.02) | 61.71 (17.52) | 62.13 (17.61) | 63.65 (7.57) | 64.02 (16.54) |
| Male gender | 11,533<br>(56.90%) | 2,385<br>(55.78%) | 6,358<br>(55.20%) | 16,396<br>(47.83%) | 19,645<br>(53.13%) |
| Female gender | 8,735<br>(43.10%) | 1,891<br>(44.22%) | 5,160<br>(44.80%) | 17,884<br>(52.17%) | 17,319<br>(46.84%) |
| Other/unknown gender | 1 (0.00%) | 0 (0.00%) | 0 (0.00%) | 0 (0.00%) | 11 (0.03%) |
| White | 11,427<br>(56.38%) | 2,492<br>(58.28%) | 6,500<br>(56.43%) | 33,166<br>(96.75%) | 14,106<br>(38.15%) |
| Asian | 2,917<br>(14.39%) | 586 (13.70%) | 1,743<br>(15.13%) | 459 (1.34%) | 767 (2.07%) |
| Black or African American | 1,089 (5.37%) | 258 (6.03%) | 590 (5.12%) | 226 (0.66%) | 5,781<br>(15.63%) |
| Other/unknown race | 4,836<br>(23.86%) | 940 (21.98%) | 2,685<br>(23.31%) | 429 (1.25%) | 16,321<br>(44.14%) |
| Hispanic | 2,647<br>(13.06%) | 539 (12.61%) | 1,374<br>(11.93%) |  | 9,623<br>(26.03%) |
| Non-hispanic | 17,622<br>(86.94%) | 3,737<br>(87.39%) | 10,144<br>(88.07%) |  | 16,105<br>(43.56%) |
| Unknown ethnicity | 0 (0.00%) | 0 (0.00%) | 0 (0.00%) |  | 11,247<br>(30.42%) |
| Sinus Rhythm | 15,587<br>(76.90%) | 3,298<br>(77.13%) | 8,940<br>(77.62%) | 33,563<br>(97.91%) |  |
| Pacemaker | 1,664 (8.21%) | 330 (7.72%) | 745 (6.47%) | 55 (0.16%) |  |
| Premature Ventricular Complexes | 1,367 (6.74%) | 287 (6.71%) | 757 (6.57%) | 1,201 (3.50%) |  |
| Left Bundle Branch Block | 975 (4.81%) | 224 (5.24%) | 535 (4.64%) | 311 (0.91%) |  |

To understand how well models generalize across sites, we additionally evaluated our models on ECGs from another healthcare system, Columbia Irving Medical Center, and a prospective population of healthy individuals, the UK BioBank cohort. The Columbia cohort consisted of 36,975 patients who received an ECG and TTE at Columbia medical center within a two week window. In that group, prevalence was similar to at Stanford (12.59%), and there were greater proportions of Black and Hispanic patients (Table 1). The UK Biobank cohort consisted of 34,280 patients from the general population who prospectively received cardiac magnetic resonance imaging, and had a much lower prevalence of LVSD, with just 96 (0.28%) cases. The population also had higher rates of normal ECGs (97.9% in the UK Biobank vs 77.6% at Stanford) and contained a greater proportion of White patients (96.75% in the UK Biobank vs 56.38% at Stanford).

### Simple models using discrete, automated measurements detect LVSD almost as well as deep learning models

The convolutional neural network trained on 12-lead ECG waveforms achieved an area under the receiver operator characteristic curve (AUROC) of 0.94 (0.93-0.94) in detecting LVSD, comparable to the 0.93 previously reported^6^ (Figure 1, Figure 2; previous work did not report a confidence interval on the computed AUC). Choosing a cutoff to balance sensitivity and specificity resulted in values of 0.86 (0.84-0.88) and 0.86 (0.86-0.87) respectively. At that cutoff it achieved a positive predictive value of 0.40 (0.37-0.42). At a sensitivity of 90%, it achieved a specificity of 0.82 (0.81-0.83). The model consisted of 159,153 trainable parameters.

**Figure 1.**
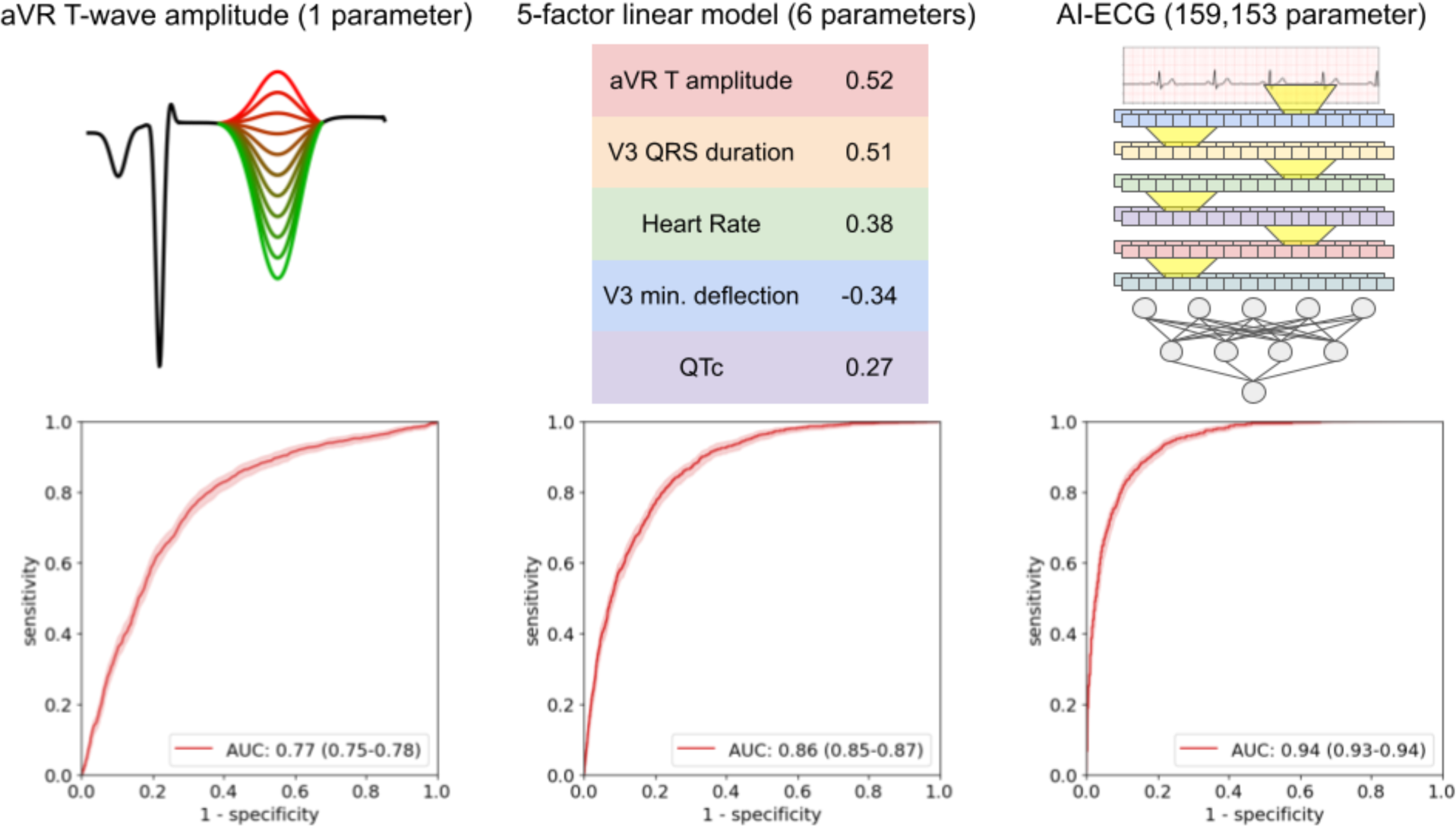
ROC curves for three risk scores for detecting LVSD. Left: the amplitude of the T-wave in lead aVR, used directly as a risk score for LVSD. Center: a linear model based on five ECG measurements. Weights based on normalized measurements are shown. Right: a deep learning model based on the ECG waveform (diagram is a simplification for illustrative purposes only).

**Figure 2.**
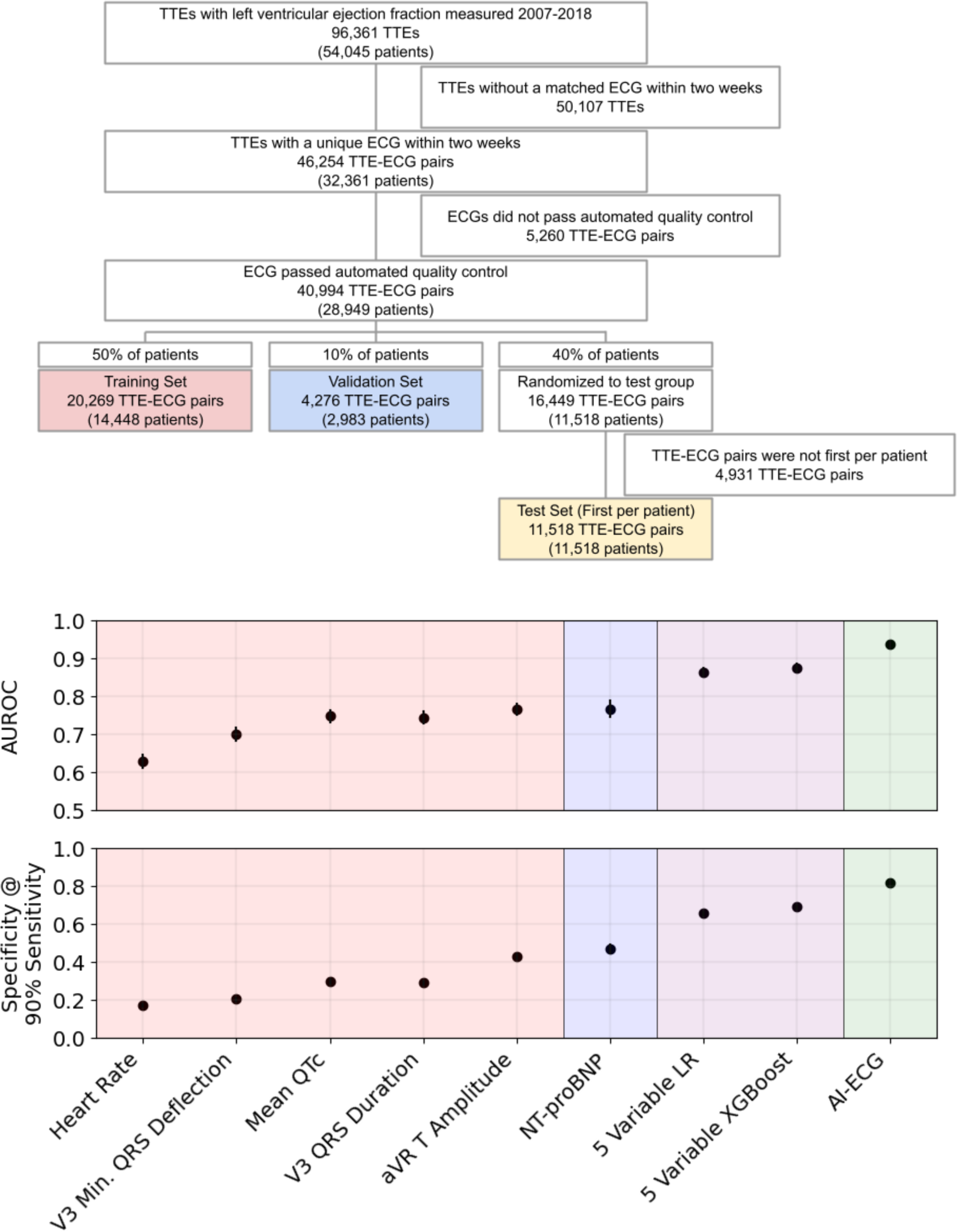
Top panel: a consort diagram for the Stanford cohort. Bottom panel: performance of several risk scores in detecting LVSD, by AUROC (area under receiver operator characteristic) and specificity at a cutoff providing 90% sensitivity. Error bars are 95% bootstrap confidence intervals.

To understand how well discrete ECG measurements can be used to detect LVSD, we next trained linear and random forest models to detect LVSD based on 555 ECG measurements extracted by the Philips TraceMaster software (listed in Supp. Table 2). Examples of such measurements (in order of increasing complexity) are the heart rate, the P wave amplitude in lead I, the area under the QRS complex in lead aVL, and the maximal T wave angle through the transverse plane. The random forest achieved an AUROC of 0.92 (0.91-0.93), not significantly different from the deep learning model (P=0.08). The best-performing random forest consisted of 50 trees of depth 7, resulting in 6,350 binary cutoffs. The linear model achieved an AUROC of 0.90 (0.89-0.91), using only 556 trainable parameters. The weights of the linear model are shown in Supp. Table 2.

Acknowledging that a 555 measurement linear model is still not easily “interpretable,” we reduced the number of measurements further, first limiting the list to familiar measurements and then using lasso regression and removing correlated features. We arrived at a shortlist of five measurements that can be easily manually assessed in a clinical setting (Table 2): the T-wave amplitude in aVR; the QRS duration in V3; the mean QTc (corrected QT interval using Bazett’s formula); the maximum negative QRS deflection in V3 (the greater of the Q and S amplitudes); and the heart rate. In all cases the correlation was positive, except for the maximum negative QRS deflection in V3 (i.e. a deeper Q or S wave in V3 indicates greater risk, while a shallower or positively inverted T wave in aVR indicates greater risk). A random forest trained on these five measurements achieved an AUC of 0.88 (0.87-0.89), while a linear model achieved an AUC of 0.86 (0.85-0.87). The random forest consisted of 20 trees of depth 4, or 80 total parameters, while the linear model used 6 trainable parameters, each one easily interpretable. Notably, the linear model achieved only slightly worse performance than the deep learning model while using only 6 versus 159,153 trainable parameters. This performance was significantly better than that of NT-proBNP (P=8.5*10^−10^), which achieved an AUROC of 0.77 (0.75-0.79; based on the 2,097 ECG-TTE pairs with an NT-proBNP measurement within 30 days). Specificity at 90% sensitivity followed a similar trend to AUC (Figure 2; Supp. Table 2).

**Table 2.** A logistic regression model for detecting LVSD. Coefficients for absolute and normed covariates are shown, along with units for each absolute covariate and P-values for each coefficient (normed and absolute P-values are the same), and AUCs for each covariate as an independent predictor.

|  | Units | Coefficient | Normed coefficient | P-Value | Independent AUC |
| --- | --- | --- | --- | --- | --- |
| aVR T Amplitude | μV | 3.69E-3 (3.35E-3 - 4.04E-3) | 0.52 (0.47 - 0.57) | 7.22E-96 | 0.77 (0.75-0.78) |
| V3 QRS Duration | ms | 2.03E-2 (1.82E-2 - 2.23E-2) | 0.51 (0.45 - 0.56) | 2.56E-81 | 0.75 (0.73-0.76) |
| Heart Rate | bpm | 1.97E-2 (1.73E-2 - 2.21E-2) | 0.38 (0.34 - 0.43) | 1.17E-58 | 0.63 (0.61-0.65) |
| V3 QRS Minimum Deflection | μV | -4.76E-4 (-5.34E-4 - -4.18E-4) | -0.34 (-0.38 - -0.30) | 7.07E-58 | 0.70 (0.68-0.72) |
| QTc | ms | 6.73E-3 (5.38E-3 - 8.08E-3) | 0.27 (0.22 - 0.33) | 1.50E-22 | 0.75 (0.73-0.77) |
| Intercept | unitless | -9.10 (-9.66 - -8.55) | -2.61 (-2.68 - -2.55) | 3.11E-225 |  |

### Single ECG measurements detect LVSD as well as NT-proBNP

For each of the 555 numerical measurements taken by the Philips TraceMaster software, we calculated the AUROC when using the measurement as an independent predictor of LVSD (Supp. Table 2). For each of the five measurements used in the small linear model, we evaluated their independent performance in detecting LVSD in the test set (Figure 2). The best-performing measurement was the T-wave amplitude in aVR, which independently achieved an AUROC of 0.77 (0.76-0.78), the same as NT-proBNP. The QRS duration in V3 and Mean QTc were also similar to NT-proBNP, achieving AUROCs of 0.75 (0.73-0.76) and 0.75 (0.74-0.77) respectively. When comparing at a 90% sensitivity cutoff however, those three ECG measurements performed significantly worse than NT-proBNP in their specificity (P=2*10^−60^-4*10^−4^).

### Simpler models perform better across sites

To understand the ability of simple and deep learning models to generalize across sites, we evaluated our models in two external cohorts, the UK Biobank cohort and Columbia cohorts. The deep learning model did not perform as well on UK Biobank data, with an AUROC of 0.74 (0.69-0.78; Table 3), but achieved good performance in the Columbia cohort, with an AUROC of 0.88 (0.87-0.88). The large difference in the UK Biobank cohort may be due to subtle differences in vendor waveform preprocessing, though we were unable to detect any differences through inspection (differences in the population or due to use of cMRI versus echocardiogram are also a plausible explanation, but are mostly ruled out by the following results). The simpler, measurement-based models, on the other hand, performed similarly to Stanford: the linear model achieved AUROCs of 0.83 (0.78-0.87) and 0.80 (0.80-0.81) in the UK Biobank and Columbia cohorts respectively, versus 0.86 at Stanford, and the random forest model achieved AUROCs of 0.82 (0.77-0.87) and 0.81 (0.80-0.82), respectively, versus 0.88 at Stanford, demonstrating the ability to generalize to radically different populations like the one in the UK Biobank cohort. The T-wave amplitude in aVR achieved similar performance in both the UK Biobank, with an AUROC of 0.78 (0.73-0.83), and Columbia, with an AUROC of 0.74 (0.74-0.75). Other Individual measurements were similarly predictive in the UK Biobank and Columbia datasets (supp. figure 2). Due to a lack of available measurements, we were unable to evaluate the 555 measurement models at external sites.

**Table 3.**
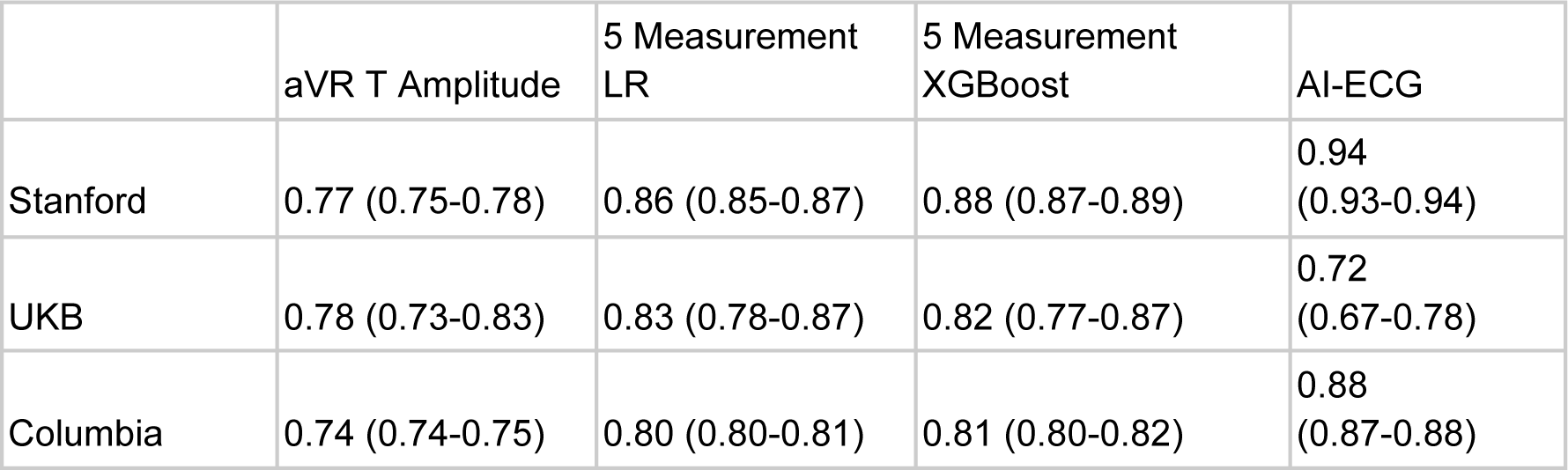
AUROCs of different predictors/models for LVSD across multiple sites.

|  | aVR T Amplitude | 5 Measurement<br>LR | 5 Measurement<br>XGBoost | AI-ECG |
| --- | --- | --- | --- | --- |
| Stanford | 0.77 (0.75-0.78) | 0.86 (0.85-0.87) | 0.88 (0.87-0.89) | 0.94<br>(0.93-0.94) |
| UKB | 0.78 (0.73-0.83) | 0.83 (0.78-0.87) | 0.82 (0.77-0.87) | 0.72<br>(0.67-0.78) |
| Columbia | 0.74 (0.74-0.75) | 0.80 (0.80-0.81) | 0.81 (0.80-0.82) | 0.88<br>(0.87-0.88) |

## Discussion

We found that simple models based on discrete, automated ECG measurements detect LVSD with impressive performance, almost as well as deep learning models using waveforms and much better than standard laboratory tests. The first strength of this study is that it is among the first to use a deep learning model using ECG waveforms, considered the optimal strategy, to benchmark the performance of simpler ECG models, revealing simple strategies that perform nearly as well as the best-performing complex models. The second strength is that it presents tangible tools that could be easier to deploy than those deep learning models. The third is that it demonstrates for the first time that these tools generalize better to different sites and populations.

While in an idealized setting medical systems would use tools with the highest accuracy possible, using simpler models has a number of benefits with respect to real-world application. As highlighted by our multicenter validation results, models with simpler inputs often exhibit stronger performance when transferred to other sites with different vendors and demographics. They are also easier to troubleshoot, and to detect unintended domain shifts in input data, since the distribution of input measurements is much simpler. These simpler models are also much more interpretable and can grant insight to physicians in ways that deep learning and even more complicated linear and tree based methods cannot. In this work, we show a continuum of models (Figures 1 and 3) which trade off complexity and performance. Notably, models based on automated measurements have been enabled by the same big data revolution that has enabled deep learning methods; previously, large datasets of automated measurements weren’t available, and the measurements were not available in real time for inference.

The five-variable linear model we trained can be directly interpreted and linked to known electrophysiologic consequences of LVSD. The development of LVSD is marked by the progressive accumulation of depolarization and repolarization abnormalities, such as abnormal QRS complexes, delays in repolarization (with prolonged QT), and more prominent T wave abnormalities, as captured by our model. Elevated heart rate^37^ and prolonged QT interval^38,39^ are both well-known to be related to severity and prognosis of LVSD. Progressive LVSD leads to decreased stroke volume and an elevated heart rate is frequently a compensatory mechanism to maintain cardiac output in addition to being a marker of atrial arrhythmias which frequently accompany heart failure. The highest weighted measurement in the regression is the T-wave amplitude in aVR, which also independently predicts LVSD with an AUROC of 0.77. This measurement was previously shown to be a strong predictor of cardiovascular and all-cause mortality^40^, despite evidence that clinicians often ignore lead aVR completely when reading ECGs^41^. An upward-facing T-wave in aVR is also correlated with ischemic etiology of cardiomyopathy^42^. Deep Q or S waves in V3 are indicative of late QRS transition which has previously been associated with risk of sudden cardiac death^43^, while prolonged QRS complexes are known to be associated with LVSD^44^. The success of the small models we present both confirms previous trends in the literature and finds new connections between the ECG and LVSD, while also providing a new and simple diagnostic tool.

Our work has limitations. While we present strong, simple models for detecting LVSD, they do not perform as well as deep learning models in terms of accuracy, but rather present different points on the continuum between complexity and performance. We used NT-proBNP as a baseline since it is the common screening tool which most closely predicts LVSD, but cases of well-compensated heart failure with low ejection fraction would not be captured by increased NT-proBNP levels. Our conclusions about LVSD likely do not transfer to all phenotypes; for example in the case of detecting atrial fibrillation in sinus rhythm, previous studies suggest deep learning achieves much higher performance^10,33^. There are also many cases where deep learning is easier to deploy or much more accurate than measurement-based methods, like when measurements are unavailable or when only a single lead is available, for example on smartwatches and other mobile devices. Our conclusion about the importance of using ECG measurements as a baseline for modeling is not as easily applied to other domains, where there are fewer high-quality interpretable features - in the specific case of ECG analysis, the available derived measurements (especially more complicated measurements like the QTc and electrical axis) are a result of over a hundred years of domain knowledge to find the optimal engineered features, which other applications have not benefited from. Finally, our work is limited to the populations which we describe, and accuracy might be diminished in different populations, although we have the benefit of working with three diverse populations (two tertiary care centers in the United States and one biobank in the United Kingdom).

We present here a set of simple methods to detect LVSD from the ECG, with performances ranging between those of standard laboratory tests and state of the art deep learning methods. In many cases, simpler methods with slightly lower accuracy based on discrete features may be better to deploy than more complicated, uninterpretable methods, and may yield improved insights into the underlying physiology. We believe there is benefit to presenting results of study techniques along the continuum of complexity as different health care systems may opt for employment of different models along this continuum based on resources and accessibility. In the setting of ECG interpretation, this is possible thanks to a wealth of domain knowledge about important ECG measurements.

## Model and code availability

For normalized inputs, the small linear model weights are shown in figure 1, and the large linear model weights in supplementary table 1. XGBoost and deep learning models, and code to train models, are available on github at [will be made available before publication].

## Data availability

UK Biobank data is available through application. Data from Stanford and Columbia Irving Medical Centers cannot be shared due to patient privacy constraints.

