## Supplemental tables and figures for "Simple Models Versus Deep Learning in Detecting Low Ejection Fraction From The Electrocardiogram"

#### Supplemental Table 1

AUC as independent predictor and LR coefficient for all ECG measurements.

| | AUC | Coefficients |
|---|---|---|
| I_tptparea | 0.78 | -0.05 |
| I_tarea | 0.77 | -0.14 |
| I_tamp | 0.77 | -0.24 |
| aVR_tptparea | 0.77 | 0.05 |
| aVR_tamp | 0.77 | 0.2 |
| V3_qrsarea | 0.76 | 0.01 |
| aVR_tarea | 0.76 | 0.02 |
| meanqtc | 0.75 | 0.25 |
| V4_qrsdur | 0.74 | -0.29 |
| V5_qrsdur | 0.74 | -0.8 |
| meanqrsdur | 0.74 | -0.33 |
| V3_qrsdur | 0.73 | 4.45 |
| V6_tptparea | 0.73 | 0.12 |
| V6_tamp | 0.73 | -0.18 |
| V4_qrsarea | 0.73 | 0.21 |
| V6_qrsdur | 0.72 | -1.59 |
| II_qrsdur | 0.72 | 1.47 |

| | | |
|---|---|---|
| V6_tarea | 0.72 | 0.08 |
| aVL_tptparea | 0.72 | 0.27 |
| V2_qrsdur | 0.72 | 0.54 |
| III_qrsdur | 0.72 | 0.38 |
| aVL_tarea | 0.72 | 0.23 |
| aVR_qrsppk | 0.72 | -0.53 |
| II_ramp | 0.71 | 0.36 |
| V4_ramp | 0.71 | 0.86 |
| aVL_tamp | 0.71 | -0.6 |
| aVF_qrsdur | 0.71 | 0.31 |
| I_qrsdur | 0.71 | 0.86 |
| tfrontaxis | 0.71 | 0.12 |
| aVL_qrsdur | 0.71 | 0.34 |
| fronttmaxangle | 0.7 | 0.08 |
| aVR_qrsdur | 0.69 | 0.28 |
| fronttinitangle | 0.69 | 0.06 |
| II_tptparea | 0.69 | 0.07 |
| I_stend | 0.69 | 0.19 |
| II_tamp | 0.69 | 0 |
| aVL_vat | 0.69 | 0.26 |
| thorizaxis | 0.69 | 0.04 |
| V2_qrsarea | 0.69 | 0 |

| | | |
|---|---|---|
| II_tarea | 0.69 | 0.1 |
| V1_qrsdur | 0.68 | -0.53 |
| V3_ramp | 0.68 | -0.31 |
| V3_sdur | 0.68 | -4.87 |
| fronttermangle | 0.68 | 0.03 |
| V3_rdur | 0.67 | -4.26 |
| V5_tamp | 0.67 | -0.11 |
| V5_tptparea | 0.67 | 0.06 |
| aVR_stend | 0.67 | 0.66 |
| frontttermmag | 0.67 | -0.42 |
| V6_stend | 0.67 | -0.72 |
| V5_ramp | 0.67 | -1.03 |
| V5_tarea | 0.67 | -0.04 |
| I_st80 | 0.67 | -0.38 |
| V6_ston | 0.67 | 0.34 |
| sagqrsmaxmag | 0.66 | 0.36 |
| aVL_stend | 0.66 | -2.22 |
| I_vat | 0.66 | -0.1 |
| I_ston | 0.66 | -1.16 |
| aVR_ston | 0.66 | -0.11 |
| I_stmid | 0.66 | 1.62 |
| frontqrstermmag | 0.65 | 0.26 |

| | | |
|---|---|---|
| transqrstermmag | 0.65 | -0.08 |
| aVF_qrsarea | 0.65 | 0.14 |
| fronttmaxmag | 0.65 | -0.36 |
| V3_vat | 0.65 | 0.09 |
| II_qrsarea | 0.65 | -0.16 |
| aVR_tdur | 0.65 | 1.31 |
| V6_st80 | 0.65 | -0.09 |
| meanrrint | 0.65 | -0.29 |
| V1_tamp | 0.65 | 0.21 |
| transqrsinitangle | 0.65 | 0.18 |
| aVL_st80 | 0.65 | 4.06 |
| transqrsmaxangle | 0.65 | 0.13 |
| aVF_ramp | 0.65 | -0.96 |
| V1_tptparea | 0.65 | -0.12 |
| V4_sdur | 0.65 | 0.4 |
| V6_stmid | 0.65 | 0.33 |
| I_stslope | 0.65 | -0.08 |
| sagqrsmaxangle | 0.64 | 0.07 |
| aVL_stslope | 0.64 | 0.32 |

| | | |
|---|---|---|
| I_rdur | 0.64 | -0.65 |
| stfrontaxis | 0.64 | -0.06 |
| aVR_st80 | 0.64 | -0.45 |
| transqrscwrot | 0.64 | 0.25 |
| V1_tarea | 0.64 | -0.12 |
| aVL_rdur | 0.64 | -0.54 |
| sagqrstermmag | 0.63 | -0.04 |
| aVR_tptpdur | 0.63 | -1.44 |
| aVR_stmid | 0.63 | -0.22 |
| lowventrate | 0.63 | -0.6 |
| II_qrsppk | 0.63 | -0.67 |
| meanventrate | 0.63 | 0.01 |
| aVL_stmid | 0.63 | -1.43 |
| I_tdur | 0.63 | 2.54 |
| III_qrsarea | 0.63 | 0.11 |
| aVL_stdur | 0.63 | -0.09 |
| V6_rdur | 0.63 | 1.4 |
| III_stdur | 0.63 | 0.25 |
| aVF_stdur | 0.63 | -0.36 |
| V6_stdur | 0.63 | -0.12 |
| V5_stdur | 0.63 | -3.98 |
| V1_stdur | 0.63 | 1.7 |

| | | |
|---|---|---|
| I_stdur | 0.63 | 0.42 |
| V3_stdur | 0.63 | 0.97 |
| fronttinitmag | 0.63 | 0.45 |
| V4_stdur | 0.63 | 1.41 |
| aVR_stdur | 0.63 | 0.11 |
| II_stdur | 0.63 | -0.41 |
| V2_stdur | 0.63 | -0.09 |
| I_ramp | 0.63 | -0.32 |
| numberofcomplexes | 0.62 | 0.35 |
| highventrate | 0.62 | 0.46 |
| II_ston | 0.62 | -0.57 |
| sthorizaxis | 0.62 | -0.01 |
| transtinitangle | 0.62 | 0.06 |
| V2_rdur | 0.62 | -0.41 |
| V2_ramp | 0.62 | -0.01 |
| V5_qrsarea | 0.62 | 0.23 |
| meanqtseg | 0.61 | -0.76 |
| ventratestddev | 0.61 | -0.27 |
| V1_ston | 0.61 | -0.24 |
| V3_samp | 0.61 | 0.3 |
| I_qrsppk | 0.61 | 0.36 |
| II_tdur | 0.61 | -0.16 |

| | | |
|---|---|---|
| II_stend | 0.61 | 2.1 |
| i40horizaxis | 0.61 | 0.02 |
| V2_sdur | 0.61 | -0.19 |
| V5_stend | 0.61 | 0.5 |
| V6_ramp | 0.61 | -1.18 |
| sagqrsinitmag | 0.61 | 0.1 |
| V1_qrsarea | 0.61 | 0.05 |
| I_tptpdur | 0.61 | -2.61 |
| sagqrsinitangle | 0.61 | 0.06 |
| III_stslope | 0.6 | 0.49 |
| sagtmaxmag | 0.6 | -0.06 |
| V1_ppppdur | 0.6 | -0.01 |
| aVR_stslope | 0.6 | 0.02 |
| V6_vat | 0.6 | 0.02 |
| V5_ston | 0.6 | -0.91 |
| transpcwrot | 0.6 | -0.03 |
| aVR_qrsarea | 0.6 | 0.01 |
| II_tptpdur | 0.6 | 0.06 |
| sagptermangle | 0.6 | -0.12 |
| V1_qdur | 0.6 | 0.57 |
| III_tamp | 0.6 | -0.07 |
| V6_stslope | 0.6 | 0.34 |

| | | |
|---|---|---|
| V2_vat | 0.6 | 0.03 |
| V2_pppparea | 0.6 | 0.04 |
| aVL_ston | 0.6 | -0.39 |
| III_tptparea | 0.6 | -0.01 |
| III_tarea | 0.6 | 0 |
| V4_rdur | 0.59 | 0.19 |
| V4_samp | 0.59 | -0.84 |
| V6_tdur | 0.59 | 0.81 |
| V1_qamp | 0.59 | -0.03 |
| V3_pppparea | 0.59 | 0.08 |
| V3_qrsppk | 0.59 | 0.39 |
| transttermmag | 0.59 | 0.47 |
| III_stend | 0.59 | -1.79 |
| III_ramp | 0.59 | -0.05 |
| V4_vat | 0.59 | 0.04 |
| aVL_qrsarea | 0.59 | 0 |
| V5_st80 | 0.59 | 0.11 |
| aVF_sdur | 0.59 | -0.35 |
| V4_pppparea | 0.59 | -0.03 |
| qtintdispersion | 0.59 | 0.05 |
| III_st80 | 0.59 | 2.41 |
| V5_tdur | 0.58 | 0.43 |

| | | |
|---|---|---|
| V5_sdur | 0.58 | 0.96 |
| V1_print | 0.58 | 0.04 |
| V5_stmid | 0.58 | 0.29 |
| V5_pamp | 0.58 | -0.08 |
| frontpinitmag | 0.58 | -0.2 |
| V3_parea | 0.58 | 0.05 |
| II_sdur | 0.58 | -1.28 |
| II_st80 | 0.58 | -1.15 |
| V2_qrsppk | 0.58 | 0.23 |
| V4_pamp | 0.58 | 0 |
| V4_parea | 0.58 | 0.01 |
| V1_stend | 0.58 | 0.88 |
| transptermangle | 0.58 | -0.04 |
| frontptermangle | 0.58 | -0.03 |
| V5_qrsppk | 0.58 | 1.21 |
| aVR_samp | 0.58 | -0.01 |
| V4_tamp | 0.58 | 0.03 |
| transtmaxangle | 0.58 | -0.11 |
| transttermangle | 0.58 | -0.01 |
| V4_tptparea | 0.58 | 0.08 |
| V3_pamp | 0.58 | -0.1 |
| II_stmid | 0.58 | -0.25 |

| | | |
|---|---|---|
| aVR_qdur | 0.58 | -0.48 |
| V3_ston | 0.58 | 0.38 |
| V4_tarea | 0.58 | 0.1 |
| V5_parea | 0.58 | 0.02 |
| V2_parea | 0.57 | 0.04 |
| V5_pppparea | 0.57 | 0.01 |
| V2_pamp | 0.57 | -0.11 |
| V2_qdur | 0.57 | -0.32 |
| V2_qamp | 0.57 | 0.07 |
| aVR_rdur | 0.57 | -0.33 |
| sagpcwrot | 0.57 | 0.01 |
| frontqrstermangle | 0.57 | -0.03 |
| III_stmid | 0.57 | -0.07 |
| III_sdur | 0.57 | -0.25 |
| V1_st80 | 0.57 | -0.23 |
| avgpcount | 0.57 | 0 |
| transpinitmag | 0.57 | -0.13 |
| sagqrstermangle | 0.57 | 0.04 |
| II_pamp | 0.57 | -0.04 |
| aVF_tptparea | 0.57 | -0.19 |
| transqrsinitmag | 0.57 | -0.4 |

| | | |
|---|---|---|
| aVL_samp | 0.57 | 0.46 |
| V4_tdur | 0.57 | -0.49 |
| sagpinitangle | 0.57 | -0.04 |
| frontpmaxmag | 0.57 | 0.15 |
| flutterfibcount | 0.57 | -0.03 |
| aVF_tarea | 0.57 | -0.12 |
| aVF_tamp | 0.57 | -0.11 |
| atrialratestddev | 0.57 | 0.03 |
| t40frontaxis | 0.57 | -0.03 |
| V1_ramp | 0.57 | 0.19 |
| V6_pamp | 0.56 | 0 |
| V2_qtint | 0.56 | 0.09 |
| V1_pppparea | 0.56 | -0.11 |
| atrialrate | 0.56 | 0.1 |
| V1_stmid | 0.56 | -0.16 |
| aVF_ston | 0.56 | 0.48 |
| V2_ppppdur | 0.56 | -0.03 |
| V6_tptpdur | 0.56 | -0.86 |
| V5_tptpdur | 0.56 | -0.42 |
| V1_samp | 0.56 | -0.2 |
| I_pppparea | 0.56 | -0.04 |
| I_parea | 0.56 | -0.04 |

| | | |
|---|---|---|
| V6_parea | 0.56 | 0.01 |
| V2_ston | 0.56 | -0.02 |
| II_prseg | 0.56 | 0.01 |
| qrsfrontaxis | 0.56 | 0.08 |
| highprint | 0.56 | 0.2 |
| frontpmaxangle | 0.56 | -0.01 |
| V6_pppparea | 0.56 | 0.01 |
| sagqrscwrot | 0.56 | 0.03 |
| V3_stmid | 0.56 | -0.54 |
| phorizaxis | 0.56 | 0.01 |
| V5_stslope | 0.56 | -0.2 |
| transtcwrot | 0.56 | 0.05 |
| transpmaxangle | 0.56 | 0.16 |
| aVR_prseg | 0.56 | 0.01 |
| aVR_pamp | 0.56 | 0.1 |
| II_print | 0.56 | 0.07 |
| II_parea | 0.56 | 0 |
| sagttermmag | 0.56 | -0.1 |
| II_pppparea | 0.56 | -0.02 |
| frontpcwrot | 0.56 | -0.04 |
| V3_qdur | 0.56 | -5.11 |

| | | |
|---|---|---|
| comppausecount | 0.56 | 0.1 |
| V3_qamp | 0.56 | 0.27 |
| aVR_qamp | 0.56 | -0.16 |
| aVR_parea | 0.56 | -0.1 |
| sagpmaxangle | 0.56 | 0.06 |
| V3_st80 | 0.56 | -0.27 |
| aVR_pppparea | 0.56 | -0.05 |
| I_prseg | 0.56 | -0.04 |
| aVF_samp | 0.56 | 0.15 |
| V1_pdur | 0.55 | 0.04 |
| V1_vat | 0.55 | -0.09 |
| fronttcwrot | 0.55 | 0.03 |
| V5_rdur | 0.55 | 0.6 |
| V6_qrsppk | 0.55 | 1.11 |
| I_samp | 0.55 | 0.19 |
| III_qtint | 0.55 | -0.18 |
| V5_samp | 0.55 | 0.71 |
| II_pdur | 0.55 | 0.06 |
| aVR_vat | 0.55 | 0.01 |
| V4_tptpdur | 0.55 | 0.43 |
| frontptermmag | 0.55 | -0.13 |
| V3_qtint | 0.55 | 0.1 |

| | | |
|---|---|---|
| transpmaxmag | 0.55 | 0.29 |
| II_samp | 0.55 | 0.04 |
| aVF_prseg | 0.55 | 0.01 |
| aVL_sdur | 0.55 | -0.47 |
| V3_pdur | 0.55 | 0.06 |
| aVL_prseg | 0.55 | 0.11 |
| aVR_sdur | 0.55 | -0.24 |
| sagpinitmag | 0.55 | 0.23 |
| aVF_tptpdur | 0.55 | 0.3 |
| III_print | 0.55 | -0.04 |
| aVL_print | 0.55 | -0.1 |
| I_pdur | 0.55 | 0 |
| aVF_print | 0.55 | -0.02 |
| V6_qamp | 0.55 | 0.16 |
| qrshorizaxis | 0.55 | -0.01 |
| meanqtint | 0.55 | 0.83 |
| aVF_pamp | 0.55 | 0.16 |
| frontpinitangle | 0.55 | 0.05 |
| V5_pdur | 0.54 | -0.1 |
| aVR_pdur | 0.54 | 0 |
| aVF_stslope | 0.54 | -0.21 |
| frontqrscwrot | 0.54 | -0.05 |

| | | |
|---|---|---|
| aVF_parea | 0.54 | -0.05 |
| frontqrsinitmag | 0.54 | 0.19 |
| sagptermmag | 0.54 | 0.29 |
| V3_prseg | 0.54 | 0.18 |
| aVF_pppparea | 0.54 | -0.05 |
| aVF_rdur | 0.54 | -0.28 |
| V6_sdur | 0.54 | 1.57 |
| V3_stend | 0.54 | 0.49 |
| aVF_tdur | 0.54 | -0.32 |
| aVL_qtint | 0.54 | 0.08 |
| V1_qrsppk | 0.54 | -0.17 |
| V1_rdur | 0.54 | 0.48 |
| V1_qtint | 0.54 | 0.06 |
| aVR_print | 0.54 | 0 |
| V1_tdur | 0.54 | -0.2 |
| transtmaxmag | 0.54 | -0.52 |
| V6_prseg | 0.54 | -0.1 |
| frontqrsinitangle | 0.54 | -0.1 |
| V6_qdur | 0.54 | 1.23 |
| V1_rpamp | 0.54 | 0.21 |
| I_qamp | 0.54 | 0.17 |
| V1_rpdur | 0.54 | 0.29 |

| | | |
|---|---|---|
| I_sdur | 0.54 | -0.65 |
| V1_prseg | 0.54 | 0.05 |
| transpinitangle | 0.54 | 0.09 |
| aVR_ramp | 0.54 | 0.1 |
| aVR_rpamp | 0.54 | 0.09 |
| V1_tpamp | 0.54 | 0.14 |
| III_qrsppk | 0.54 | 0.57 |
| V6_pdur | 0.54 | -0.03 |
| I_pamp | 0.54 | 0.15 |
| V1_tparea | 0.54 | -0.06 |
| V3_tdur | 0.54 | -0.49 |
| V1_stslope | 0.53 | -0.13 |
| V1_tpdur | 0.53 | -0.1 |
| V4_qtint | 0.53 | -0.06 |
| t40horizaxis | 0.53 | -0.09 |
| V4_pdur | 0.53 | -0.02 |
| V3_print | 0.53 | -0.15 |
| aVR_rpdur | 0.53 | -0.22 |
| V5_vat | 0.53 | 0.01 |
| V1_parea | 0.53 | 0 |
| V2_print | 0.53 | 0 |
| V6_samp | 0.53 | 0.38 |

| | | |
|---|---|---|
| V5_prseg | 0.53 | 0.03 |
| meanprint | 0.53 | -0.12 |
| V2_samp | 0.53 | 0.16 |
| III_prseg | 0.53 | -0.01 |
| V6_print | 0.53 | 0.04 |
| V5_print | 0.53 | -0.01 |
| aVF_vat | 0.53 | 0.04 |
| I_print | 0.53 | 0.02 |
| II_stslope | 0.53 | -0.08 |
| V1_pamp | 0.53 | 0.05 |
| III_samp | 0.53 | 0.42 |
| aVL_parea | 0.53 | -0.01 |
| V4_qrsppk | 0.53 | -0.98 |
| sagpmaxmag | 0.53 | -0.49 |
| pfrontaxis | 0.53 | -0.04 |
| aVL_pppparea | 0.53 | -0.03 |
| V4_qdur | 0.53 | 0.32 |
| V2_stmid | 0.53 | 0.08 |
| V4_prseg | 0.53 | 0 |
| III_tdur | 0.53 | -0.06 |
| aVF_pdur | 0.53 | -0.01 |
| transqrsmaxmag | 0.53 | 0.31 |

| | | |
|---|---|---|
| V4_qamp | 0.53 | -0.7 |
| V3_tptpdur | 0.53 | 0.42 |
| printstddev | 0.53 | -0.13 |
| V2_st80 | 0.53 | 0.07 |
| V2_tdur | 0.53 | 1.78 |
| III_vat | 0.52 | 0.11 |
| aVL_qamp | 0.52 | 0.53 |
| III_qdur | 0.52 | -0.44 |
| V4_print | 0.52 | 0 |
| III_ppppdur | 0.52 | -0.07 |
| III_rpamp | 0.52 | 0.01 |
| V6_tpdur | 0.52 | 0.59 |
| aVF_stend | 0.52 | -1.27 |
| V1_ppamp | 0.52 | 0.25 |
| V1_pparea | 0.52 | -0.31 |
| III_rpdur | 0.52 | -0.36 |
| V5_qtint | 0.52 | -0.01 |
| aVF_qtint | 0.52 | 0.07 |
| III_pparea | 0.52 | 0.07 |
| III_tptpdur | 0.52 | 0.09 |
| aVL_rpdur | 0.52 | -0.23 |
| aVL_rpamp | 0.52 | -0.17 |

| | | |
|---|---|---|
| V6_qrsarea | 0.52 | 0.11 |
| I_tpdur | 0.52 | 1.88 |
| I_ppppdur | 0.52 | 0.03 |
| aVL_tdur | 0.52 | 0.62 |
| I_rpdur | 0.52 | -0.2 |
| I_rpamp | 0.52 | -0.02 |
| III_ppamp | 0.52 | -0.1 |
| V5_tpdur | 0.52 | 0.24 |
| aVL_pamp | 0.52 | 0.01 |
| aVR_qtint | 0.52 | 0.17 |
| I_qdur | 0.52 | -0.4 |
| sagttermangle | 0.52 | 0 |
| aVL_tptpdur | 0.52 | -0.71 |
| V3_stslope | 0.52 | -0.03 |
| aVL_ppppdur | 0.52 | 0.11 |
| III_pamp | 0.52 | -0.13 |
| III_qamp | 0.52 | 0.19 |
| meanprseg | 0.52 | -0.09 |
| V2_rpdur | 0.52 | -0.41 |
| V2_rpamp | 0.52 | 0.07 |
| III_ston | 0.52 | -0.31 |
| III_rdur | 0.52 | -0.42 |

| | | |
|---|---|---|
| II_vat | 0.52 | 0.03 |
| V4_tpdur | 0.52 | -0.31 |
| V2_tptpdur | 0.52 | -1.69 |
| V6_ppppdur | 0.52 | 0 |
| V5_qamp | 0.51 | 0.5 |
| III_parea | 0.51 | 0.03 |
| V4_stend | 0.51 | -0.45 |
| aVF_ppamp | 0.51 | -0.04 |
| aVR_tpdur | 0.51 | 0.94 |
| III_pppparea | 0.51 | 0.09 |
| aVF_pparea | 0.51 | 0.04 |
| I_qrsarea | 0.51 | 0.13 |
| V1_tptpdur | 0.51 | 0.02 |
| II_qamp | 0.51 | 0.08 |
| V2_stend | 0.51 | -0.16 |
| V3_ppppdur | 0.51 | 0.01 |
| V2_prseg | 0.51 | -0.07 |
| notavgpbeats | 0.51 | -0.03 |
| aVF_qdur | 0.51 | -0.19 |
| V5_qdur | 0.51 | 0.91 |
| aVF_stmid | 0.51 | -0.75 |
| V4_stmid | 0.51 | -0.16 |

| | | |
|---|---|---|
| V6_tpamp | 0.51 | -0.09 |
| I_qtint | 0.51 | 0.05 |
| lowprint | 0.51 | 0.04 |
| V6_tparea | 0.51 | 0.11 |
| deltawavecount | 0.51 | 0.04 |
| II_tpdur | 0.51 | -0.15 |
| V6_qtint | 0.51 | -0.04 |
| V5_ppppdur | 0.51 | -0.01 |
| aVF_st80 | 0.51 | 1.49 |
| V2_tamp | 0.51 | 0.16 |
| aVR_ppppdur | 0.51 | 0 |
| V4_spamp | 0.51 | -0.06 |
| V4_spdur | 0.51 | 0.1 |
| V5_rpdur | 0.51 | 0.2 |
| V5_rpamp | 0.51 | -0.11 |
| V3_tamp | 0.51 | 0.08 |
| III_pdur | 0.51 | 0.07 |
| V4_rpamp | 0.51 | 0.17 |
| V4_ppppdur | 0.51 | 0.04 |
| aVR_tpamp | 0.51 | 0.1 |
| V4_rpdur | 0.51 | -0.01 |
| transtinitmag | 0.51 | -0.21 |

| | | |
|---|---|---|
| II_tpamp | 0.51 | 0.2 |
| II_ppamp | 0.51 | 0 |
| transqrstermangle | 0.51 | 0.04 |
| V5_ppamp | 0.51 | 0.06 |
| V2_stslope | 0.51 | -0.08 |
| aVL_pdur | 0.51 | -0.03 |
| V5_spdur | 0.51 | 0.11 |
| V5_spamp | 0.51 | 0.04 |
| II_rpdur | 0.51 | -0.34 |
| II_rpamp | 0.51 | 0.06 |
| aVF_qrsppk | 0.51 | 0.6 |
| II_spdur | 0.51 | -0.35 |
| II_spamp | 0.51 | -0.07 |
| II_tparea | 0.51 | -0.12 |
| aVR_tparea | 0.51 | -0.01 |
| V5_pparea | 0.51 | -0.07 |
| aVL_spdur | 0.51 | -0.05 |
| aVL_spamp | 0.51 | 0.02 |
| I_tpamp | 0.51 | -0.23 |
| I_spdur | 0.51 | -0.02 |
| II_qdur | 0.51 | -1.19 |
| I_spamp | 0.51 | 0.1 |

| | | |
|---|---|---|
| V4_stslope | 0.51 | -0.11 |
| aVF_qamp | 0.51 | 0.21 |
| V3_tpdur | 0.51 | -0.32 |
| V3_ppamp | 0.51 | 0.03 |
| V6_pparea | 0.51 | -0.03 |
| I_tparea | 0.51 | 0.17 |
| deltawavepercent | 0.51 | -0.1 |
| aVF_tpdur | 0.51 | -0.22 |
| trigeminycount | 0.51 | 0.23 |
| trigeminystring | 0.51 | -0.24 |
| V3_tarea | 0.51 | -0.09 |
| III_tpdur | 0.51 | -0.07 |
| aVF_ppppdur | 0.51 | 0.03 |
| aVF_rpamp | 0.51 | -0.15 |
| V3_pparea | 0.51 | -0.07 |
| II_pparea | 0.51 | -0.01 |
| i40frontaxis | 0.5 | 0.1 |
| aVF_rpdur | 0.5 | -0.17 |
| V6_ppamp | 0.5 | 0 |
| V2_tpdur | 0.5 | 1.13 |
| V4_st80 | 0.5 | 0.59 |
| V6_rpamp | 0.5 | -0.24 |

| | | |
|---|---|---|
| V6_rpdur | 0.5 | 0.37 |
| III_spdur | 0.5 | -0.03 |
| I_ppamp | 0.5 | -0.01 |
| V4_tpamp | 0.5 | 0.09 |
| III_spamp | 0.5 | 0 |
| V5_tpamp | 0.5 | 0.02 |
| V3_tptparea | 0.5 | -0.1 |
| aVL_qrsppk | 0.5 | 0.61 |
| V3_tparea | 0.5 | -0.21 |
| bigeminycount | 0.5 | 0.07 |
| bigeminystring | 0.5 | -0.13 |
| II_qtint | 0.5 | 0.15 |
| aVL_tpdur | 0.5 | 0.35 |
| V5_tparea | 0.5 | -0.1 |
| V4_tparea | 0.5 | -0.09 |
| aVL_pparea | 0.5 | 0.07 |
| V2_tparea | 0.5 | 0.01 |
| aVL_ppamp | 0.5 | -0.02 |
| V1_spamp | 0.5 | -0.04 |
| I_pparea | 0.5 | 0.02 |
| V1_spdur | 0.5 | 0.05 |
| aVR_pparea | 0.5 | -0.05 |

| | | |
|---|---|---|
| V3_tpamp | 0.5 | 0.24 |
| aVF_tparea | 0.5 | 0.03 |
| V2_spamp | 0.5 | 0.05 |
| V2_spdur | 0.5 | 0.03 |
| V2_ppamp | 0.5 | -0.12 |
| V6_spdur | 0.5 | 0.08 |
| V6_spamp | 0.5 | -0.14 |
| V2_tpamp | 0.5 | -0.02 |
| aVL_tpamp | 0.5 | -0.02 |
| aVF_tpamp | 0.5 | -0.04 |
| aVL_tparea | 0.5 | -0.09 |
| III_tparea | 0.5 | -0.08 |
| V4_ston | 0.5 | 0.15 |
| V2_pdur | 0.5 | -0.03 |
| III_tpamp | 0.5 | 0.09 |
| aVL_qdur | 0.5 | -0.2 |
| aVL_ramp | 0.5 | -0.79 |
| V4_pparea | 0.5 | 0.03 |
| V3_spamp | 0.5 | 0.06 |
| V3_spdur | 0.5 | -0.9 |
| V2_pparea | 0.5 | 0.09 |
| aVR_spamp | 0.5 | 0.05 |

| | | |
|---|---|---|
| aVR_spdur | 0.5 | -0.09 |
| V1_sdur | 0.5 | 0.5 |
| aVF_spamp | 0.5 | -0.03 |
| aVF_spdur | 0.5 | -0.14 |
| aVR_ppamp | 0.5 | 0.04 |
| sagtmaxangle | 0.5 | -0.05 |
| II_ppppdur | 0.5 | -0.03 |
| V2_tarea | 0.5 | -0.14 |
| V2_tptparea | 0.5 | -0.15 |
| V3_rpdur | 0.5 | -2.3 |
| wenckcount | 0.5 | -0.08 |
| wenckstring | 0.5 | -0.31 |
| V3_rpamp | 0.5 | -0.06 |
| II_rdur | 0.5 | -1.13 |
| V4_ppamp | 0.5 | -0.04 |

#### Supplemental Table 2

| | Stanford | Stanford | Stanford | Stanford | Stanford |
|---|---|---|---|---|---|
| | AUC | Sensitivity @ 90% | Specificity @ 90% | Sensitivity @ equal specificity | PPV |
| Heart Rate | 0.63 (0.61-0.64) | 0.17 (0.15-0.19) | 0.18 (0.17-0.18) | 0.56 (0.53-0.59) | 0.14 (0.13-0.15) |
| V3 Min. QRS Deflection | 0.70 (0.68-0.72) | 0.37 (0.34-0.39) | 0.21 (0.20-0.21) | 0.66 (0.63-0.69) | 0.17 (0.16-0.18) |
| Mean QTc | 0.75 (0.73-0.77) | 0.39 (0.36-0.42) | 0.30 (0.29-0.31) | 0.69 (0.67-0.72) | 0.20 (0.19-0.21) |
| V3 QRS Duration | 0.75 (0.73-0.76) | 0.36 (0.33-0.39) | 0.29 (0.28-0.30) | 0.69 (0.66-0.72) | 0.20 (0.19-0.21) |
| aVR T Amplitude | 0.77 (0.75-0.78) | 0.34 (0.31-0.36) | 0.43 (0.42-0.44) | 0.72 (0.69-0.74) | 0.21 (0.20-0.23) |
| NT-proBNP | 0.77 (0.74-0.79) | 0.33 (0.28-0.37) | 0.47 (0.45-0.50) | 0.70 (0.66-0.74) | 0.40 (0.36-0.43) |
| 5 Variable LR | 0.86 (0.85-0.87) | 0.57 (0.55-0.60) | 0.66 (0.65-0.67) | 0.79 (0.76-0.81) | 0.28 (0.27-0.30) |
| 5 Variable XGBoost | 0.88 (0.87-0.89) | 0.59 (0.56-0.62) | 0.69 (0.68-0.70) | 0.80 (0.77-0.82) | 0.29 (0.28-0.31) |
| 555 Variable LR | 0.90 (0.89-0.91) | 0.71 (0.68-0.73) | 0.72 (0.72-0.73) | 0.82 (0.80-0.84) | 0.33 (0.31-0.35) |
| 555 Variable XGBoost | 0.92 (0.91-0.93) | 0.76 (0.73-0.78) | 0.79 (0.78-0.80) | 0.84 (0.82-0.87) | 0.37 (0.35-0.39) |
| AI-ECG | 0.94 (0.93-0.94) | 0.80 (0.78-0.83) | 0.82 (0.81-0.83) | 0.86 (0.84-0.88) | 0.40 (0.38-0.42) |

| | UKB | UKB | UKB | UKB | UKB |
|---|---|---|---|---|---|
| | AUC | Sensitivity @ 90% | Specificity @ 90% | Sensitivity @ equal specificity | PPV |
| Heart Rate | 0.67 (0.61-0.73) | 0.42 (0.32-0.52) | 0.15 (0.15-0.16) | 0.64 (0.53-0.73) | 0.00 (0.00-0.01) |
| V3 Min. QRS Deflection | 0.59 (0.52-0.66) | 0.26 (0.17-0.35) | 0.12 (0.11-0.12) | 0.55 (0.44-0.65) | 0.00 (0.00-0.00) |

| | | | | | |
|---|---|---|---|---|---|
| Mean QTc | 0.68 (0.62-0.75) | 0.38 (0.27-0.48) | 0.13 (0.12-0.13) | 0.64 (0.54-0.73) | 0.00 (0.00-0.01) |
| V3 QRS Duration | 0.68 (0.62-0.74) | 0.38 (0.28-0.47) | 0.16 (0.16-0.17) | 0.62 (0.53-0.72) | 0.00 (0.00-0.01) |
| aVR T Amplitude | 0.78 (0.73-0.83) | 0.49 (0.39-0.60) | 0.38 (0.38-0.39) | 0.71 (0.61-0.80) | 0.01 (0.00-0.01) |
| NT-proBNP | | | | | |
| 5 Variable LR | 0.83 (0.78-0.87) | 0.60 (0.51-0.71) | 0.48 (0.47-0.48) | 0.73 (0.62-0.81) | 0.01 (0.01-0.01) |
| 5 Variable XGBoost | 0.82 (0.77-0.87) | 0.57 (0.47-0.67) | 0.44 (0.43-0.44) | 0.77 (0.67-0.85) | 0.01 (0.01-0.01) |
| 555 Variable LR | | | | | |
| 555 Variable XGBoost | | | | | |
| AI-ECG | 0.72 (0.67-0.78) | 0.34 (0.26-0.44) | 0.36 (0.35-0.36) | 0.66 (0.56-0.75) | 0.01 (0.00-0.01) |

| | Columbia | Columbia | Columbia | Columbia | Columbia |
|---|---|---|---|---|---|
| | AUC | Sensitivity @ 90% | Specificity @ 90% | Sensitivity @ equal specificity | PPV |
| Heart Rate | 0.57 (0.57-0.58) | 0.11 (0.10-0.12) | 0.09 (0.09-0.09) | 0.53 (0.52-0.55) | 0.15 (0.15-0.16) |
| V3 Min. QRS Deflection | 0.64 (0.63-0.65) | 0.29 (0.28-0.30) | 0.13 (0.12-0.13) | 0.61 (0.59-0.62) | 0.18 (0.17-0.19) |
| Mean QTc | 0.72 (0.71-0.73) | 0.35 (0.34-0.37) | 0.26 (0.25-0.26) | 0.66 (0.65-0.68) | 0.22 (0.22-0.23) |
| V3 QRS Duration | 0.64 (0.63-0.65) | 0.27 (0.26-0.28) | 0.18 (0.18-0.19) | 0.56 (0.55-0.58) | 0.18 (0.17-0.18) |
| aVR T Amplitude | 0.74 (0.74-0.75) | 0.28 (0.27-0.29) | 0.39 (0.38-0.39) | 0.69 (0.67-0.70) | 0.25 (0.24-0.26) |
| NT-proBNP | | | | | |
| 5 Variable LR | 0.80 (0.80-0.81) | 0.44 (0.42-0.45) | 0.49 (0.49-0.50) | 0.73 (0.72-0.74) | 0.28 (0.27-0.29) |
| 5 Variable XGBoost | 0.81 (0.80-0.82) | 0.41 (0.40-0.43) | 0.55 (0.54-0.55) | 0.73 (0.72-0.75) | 0.28 (0.28-0.29) |

| | | | | | |
|---|---|---|---|---|---|
| 555 Variable LR | | | | | |
| 555 Variable XGBoost | | | | | |
| AI-ECG | 0.88 (0.87-0.88) | 0.65 (0.63-0.66) | 0.66 (0.66-0.67) | 0.80 (0.79-0.81) | 0.37 (0.36-0.38) |

### Supplemental Figures

#### Supplemental Figure 1

Performance of several risk scores in detecting LVSD, by AUROC (area under receiver operator characteristic), specificity at a cutoff providing 90% sensitivity, sensitivity at a cutoff providing 90% sensitivity, sensitivity at a cutoff balancing sensitivity and specificity, and positive predictive value. Error bars are 95% bootstrap confidence intervals.

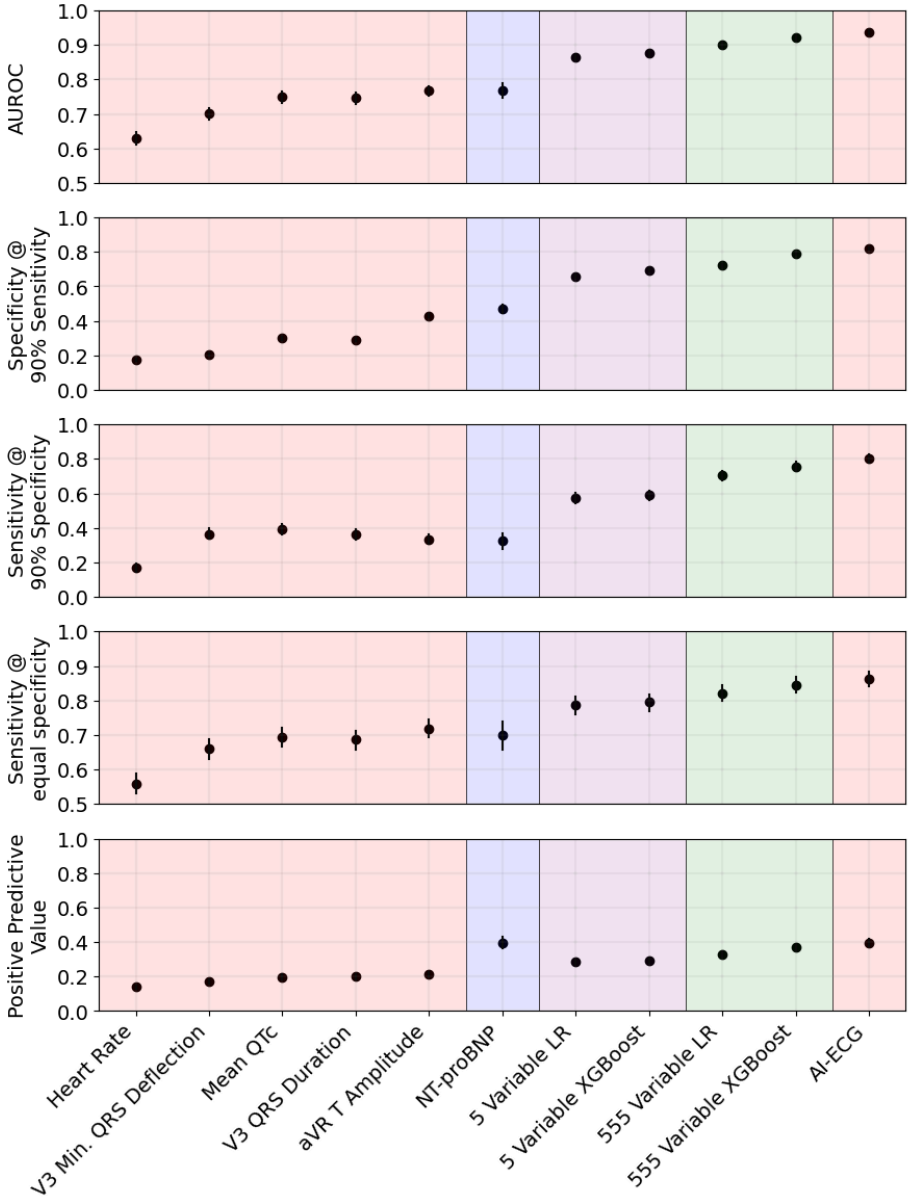
